# Ten-Year Risk of Primary Malignant Brain Tumors After Buprenorphine vs Naltrexone Exposure: A Retrospective Cohort Study Using the Epic Cosmos Database

**DOI:** 10.64898/2025.12.31.25343282

**Authors:** Kevin W. Hoffman, Rima Abram, Damisi Akinpelu, Riyad Abdalla, Jochen Meyer

**Affiliations:** School of Medicine, Baylor College of Medicine, Houston, TX, USA; Department of Neurology, Baylor College of Medicine, Houston, TX, USA

## Abstract

**Background:** Short-term neurobiological effects of opioids are well characterized, yet the long-term consequences of substance exposure on malignancy risk remain poorly understood. Ethical constraints limit prospective analysis of opioid exposure and primary malignant brain tumor (PMBT) risk in human patients, making large-scale electronic health record (EHR) analyses essential. This study evaluates the 10-year incidence of PMBTs among adults with opioid-abuse disorder related diagnoses exposed to either buprenorphine or naltrexone.

**Methods:** We conducted a retrospective cohort study using the Epic Cosmos EHR database. Cohorts were defined by exclusive exposure to buprenorphine or naltrexone, with a non-exposed comparator cohort. PMBT incidence within 1-120 months post-exposure was identified. Due to platform constraints, multivariable regression could not be performed; instead, confounding was addressed using Mantel-Haenszel stratification across age, sex, and race.

**Results:** There was no significant difference in 10-year PMBT incidence between patients exposed exclusively to buprenorphine versus naltrexone (Mantel-Haenszel OR = 1.028, 95% CI 0.968-1.092; p = 0.381), however PMBT risk was lower for opioid disorder patients receiving either drug than for the control group. Sex-specific analyses suggested women exposed to naltrexone had significantly lower PMBT incidence relative to buprenorphine exposure (OR = 0.783, 95% CI 0.617-0.994; p = 0.044), whereas men showed a non-significant increase (OR = 1.211, 95% CI 0.939-1.564; p = 0.139).

**Conclusions:** Buprenorphine and naltrexone were associated with comparable overall 10-year PMBT risk among adults with opioid-related diagnoses. The sex-specific interaction suggests naltrexone may be associated with reduced PMBT incidence in women, warranting further investigation. These findings contribute to the long-term neuro-oncologic safety profiles of opioid use disorder treatments.

**Key Points:** Buprenorphine and naltrexone were associated with comparable 10-year risks of developing primary malignant brain tumors among adults with opioid-related diagnoses.

A significant sex-specific interaction was observed, with naltrexone exposure linked to lower primary malignant brain tumor incidence in women, suggesting a potential effect modifier warranting further investigation.

Both buprenorphine and naltrexone prescription in opioid use disorder patients are associated with reduced long-term tumor incidence compared to the unexposed cohort.

## 1. Introduction

Basic and clinical research continues to establish the effects of various neuroactive substances on brain physiology. Most of these discussed effects, however, are experienced in a relatively brief timeframe: intoxication effects of opioids can peak in less than ninety minutes, and withdrawal symptom onset can begin within seventy-two hours [1]. Investigation of the longer-term effects of chronic substance use on neurons is lacking yet has great potential to shed light on risk factors for developing pathologies such as primary malignant brain tumors (PMBTs). As it is highly unethical to attempt establishing causality between opioid use and brain tumors via prospective experimental design, databases are often employed for exposure comparisons [2]. Studies have been using databases to compare cancer rates in populations exposed and unexposed to opioids for some time, but there remains a gap of knowledge in exposure-non-exposure studies in brain cancers [3]. In this retrospective cohort study of records drawn from the EPIC Cosmos EHR database, we examined the 10-year incidence of primary malignant brain tumors in patients with either buprenorphine or naltrexone exposure secondary to a diagnosed opioid-related condition.

## 2. Methods

We conducted a retrospective cohort study using the Epic Cosmos electronic health record (EHR) database, which aggregates de-identified patient data across participating health systems. Patients were included if they were ≥18 years old with an opioid-related diagnosis (ICD-10-CM F11 or T40.0-T40.3 codes) between Jan 1, 2016, and Jan 1, 2025. Patients with prior diagnoses that could confound primary brain tumor (PMBT) outcomes were excluded, including pre-existing primary brain tumors (C71, D33, D43), HIV infection (B20), intracranial injury (S06), phakomatoses (Q85), secondary brain malignancies (C79), or infectious/inflammatory central nervous system conditions (G00-G09).

Exposure cohorts were defined based on dispensed medication history of buprenorphine or naltrexone, excluding any exposure of the other medication. A non-exposure cohort was defined as patients never receiving either medication. Any patients exposed to methadone were also excluded to prevent confounding from other exposures. These cohorts were subdivided into patients who developed a PMBT (C71) and those who did not within 1-120 months following medication exposure. Baseline characteristics of each cohort included demographics (age, sex, race/ethnicity), body mass index (BMI), smoking status, and psychiatric or medical comorbidities commonly associated with substance use or brain tumor risk (alcohol use disorder F10, other substance use disorders F12-F19, hepatitis B19, type 2 diabetes E11, hypertension I15, seizure disorder G40, depression F33, bipolar disorder F31, schizophrenia F20, and anxiety disorders F41). Counts <10 were suppressed in accordance with Cosmos data use policies to preserve patient privacy.

Due to platform constraints in Cosmos SlicerDicer, regression-based multivariable adjustment was not feasible. Instead, we used Mantel-Haenszel stratification to control for confounding by age, sex, and race, consistent with prior EHR-based studies [4]. Stratum-specific relationships were explored via chi-squared and Fischer’s exact tests, with subsequent analysis confirming heterogeneity via Woolf’s tests. Our methods of analyses closely follow other similar studies and in accordance with STROBE guidelines[5].

## 3. Results

We found no significant difference in 10-year development of PMBTs between the pure Buprenorphine exposure and pure Naltrexone exposure cohorts when controlling for age, sex, race, BMI, smoking status, and comorbidities (MH, OR = 1.028, 95% CI = 0.968-1.092, p = 0.381). However, subgroup analyses identified sex as a potential effect modifier, where women saw 22% less PMBT incidence after Naltrexone exposure (OR = 0.783, 95% CI = 0.617-0.994, p = 0.044, see table 1). This compared to men who had a non-significant increase (OR = 1.211, 95% CI = 0.939-1.564, p = 0.139). Woolf’s test confirmed this interaction (chi^2 = 6.012, df = 1, p = 0.014). No other stratum was significant (table 1). Comparison of either medication with the unexposed cohort revealed a similar protective effect with buprenorphine (MH, OR = 0.920, 95% CI = 0.876-965, p < 0.001, see suppl. table 1 for unexposed patient data) and naltrexone (MH, OR = 0.944, 95% CI = 0.902-0.987, p < 0.001). This protective effect persisted across age, sex, race, smoking status, and comorbidities, but not for BMI. Specifically, underweight patients saw a significant increase in PMBT incidence following buprenorphine exposure (χ^2^, OR = 2.335, 95% CI = 1.320-4.131, p = 0.008) and naltrexone exposure (χ^2^, OR = 2.002, 95% CI = 1.179-3.399, p = 0.019).

## 4. Discussion

In this exploratory, retrospective cohort study of records drawn from the EPIC Cosmos EHR database, we examined the 10-year incidence of primary malignant brain tumors (PMBTs) in relation to patient exposure to buprenorphine vs. naltrexone in the context of diagnosed opioid use disorder-related conditions. We found no significant difference in PMBT incidence between patients exposed purely to buprenorphine and those exposed purely to naltrexone, when controlling for age, sex, and race using Mantel-Haenszel stratification. This finding suggests both pharmacological treatments for opioid dependence have comparable safety profiles not only in relation to cancer incidence, but also specific to PMBTs [3]. Additionally, both buprenorphine and naltrexone were not associated with increased PMBT risk, validating the safety of prescription opioid use disorder medications. In fact, they served similar protective effects when compared to an unexposed cohort, warranting further research into potential underlying mechanisms.

A key secondary finding from our subgroup analyses was that, among women, pure naltrexone exposure correlates with a significantly lower incidence of PMBTs than pure buprenorphine exposure, confirmed by Woolf’s test. Men displayed a non-significant increase in PMBT incidence following pure naltrexone exposure compared to pure buprenorphine exposure. This difference indicates that sex is a potential effect modifier; possible mechanisms include differences in immune modulation of metabolism, opioid receptor expression, or social determinants [6]. Future interventional animal model studies may focus on elucidating the biological mechanisms behind this sex difference.

The large-scale nature of the EHR networks facilitates our focus on rare malignancies such as PMBTs since it draws from a large sample size of over 300 million patients with geographical and demographic diversity. However, this method of research is not without its limitations. Despite stratification, we could not complete a causal analysis due to potential confounding factors. Indeed, additional studies are needed to confirm our findings comparing exposed to unexposed cohorts since platform constraints prevented line-level logistic or linear regression. Moreover, diagnostic protocols may differ between medical settings, leading to inconsistencies in classifications. Finally, there could be dose-dependent effects that we were not able to analyze due to the limitations within the Cosmos database.

In conclusion, buprenorphine and naltrexone exposure are associated with similar risks of developing PMBTs, and similar improvement compared to non-exposure. Further research is warranted to elucidate the mechanism behind the sex-specific and BMI-specific differences in incidence. Understanding the correlation between opioid exposure and malignancy risk is crucial to optimizing drug safety and effective treatments for opioid use disorder.

## Data Availability

The data supporting the findings of this study are available from the Epic COSMOS EHR database. Restrictions apply to the availability of these data.

