## Supplementary material for "Ten-Year Risk of Primary Malignant Brain Tumors After Buprenorphine vs Naltrexone Exposure: A Retrospective Cohort Study Using the Epic Cosmos Database": Table 1

|  | **Buprenorphine** | | **Buprenorphine** | | **Naltrexone** | | **Naltrexone** | | **Statistic** |
| --- | --- | --- | --- | --- | --- | --- | --- | --- | --- |
|  | **PMBT N=233** | | **None N=419,126** | | **PMBT N=297** | | **None N=485,956** | | **RR (95% CI), p** |
|  | Count | **%** | Count | **%** | Count | **%** | Count | **%** |  |
| **Age group** |  |  |  |  |  |  |  |  |  |
| 18-45 | 88 | 37.8 | 222,220 | 53.0 | 138 | 46.5 | 293,981 | 60.5 | 0.87 (0.67-1.14), p=0.31 |
| 45-65 | 90 | 38.6 | 145,364 | 34.7 | 117 | 39.4 | 141,452 | 29.1 | 0.86 (0.66-1.14), p=0.29 |
| ≥65 | 55 | 23.6 | 51,542 | 12.3 | 42 | 14.1 | 28,849 | 5.9 | 1.07 (0.72-1.60), p=0.74 |
| **Sex** |  |  |  |  |  |  |  |  |  |
| Female | 114 | 48.9 | 198,241 | 47.3 | 123 | 41.4 | 259,211 | 53.3 | 0.78 (0.62-0.99), p=0.04 |
| Male | 119 | 51.1 | 220,795 | 52.7 | 156 | 52.5 | 226,639 | 46.6 | 1.21 (0.94-1.56), p=0.14 |
| **Race** |  |  |  |  |  |  |  |  |  |
| White | 199 | 85.4 | 353,644 | 84.4 | 240 | 80.8 | 410,792 | 84.5 | 0.96 (0.80-1.16), p=0.70 |
| Black | 27 | 11.6 | 53,031 | 12.7 | 32 | 10.8 | 61,643 | 12.7 | 0.98 (0.59-1.64), p=0.94 |
| Asian | <10 |  | 3,651 | 0.9 | <10 | N/A | 4,297 | 0.9 | 1.18 (0.34-4.07), p=1.00 |
| Other | 29 | 12.4 | 58,404 | 13.9 | 38 | 12.8 | 65,931 | 13.6 | 0.87 (0.53-1.40), p=0.54 |
| **BMI** |  |  |  |  |  |  |  |  |  |
| <18.5 | 14 | 6.0 | 20,088 | 4.8 | 17 | 5.7 | 24,373 | 5.0 | 1.16 (0.57-2.35), p=0.68 |
| 18.5-24.9 | 76 | 32.6 | 151,100 | 36.1 | 94 | 31.6 | 175,773 | 36.2 | 1.03 (0.76-1.40), p=0.82 |
| 25-29.9 | 84 | 36.1 | 147,391 | 35.2 | 111 | 37.4 | 173,835 | 35.8 | 0.99 (0.74-1.31), p=0.94 |
| ≥30 | 96 | 41.2 | 131,420 | 31.4 | 111 | 37.4 | 149,416 | 30.7 | 1.11 (0.84-1.46), p=0.45 |
| **Smoking** |  |  |  |  |  |  |  |  |  |
| Never | 48 | 20.6 | 69,676 | 16.6 | 62 | 20.9 | 80,347 | 16.5 | 0.89 (0.61-1.30), p=0.56 |
| Current | 100 | 42.9 | 226,598 | 54.1 | 118 | 39.7 | 262,429 | 54.0 | 0.98 (0.75-1.28), p=0.89 |
| Former | 80 | 34.3 | 106,446 | 25.4 | 93 | 31.3 | 124,722 | 25.7 | 1.01 (0.74-1.36), p=0.96 |
| **Underlying conditions** | |  |  |  |  |  |  |  |  |
| AUD F10 | 40 | 17.2 | 86,308 | 20.6 | 56 | 18.9 | 122,321 | 25.2 | 1.01 (0.67-1.52), p=0.95 |
| SUD F12-19 | 176 | 75.5 | 332,560 | 79.3 | 214 | 72.1 | 390,809 | 80.4 | 0.97 (0.79-1.18), p=0.74 |
| Hepatitis B19 | 26 | 11.2 | 58,869 | 14.0 | 39 | 13.1 | 70,147 | 14.4 | 0.79 (0.48-1.30), p=0.36 |
| T2DM E11 | 58 | 24.9 | 63,651 | 15.2 | 67 | 22.6 | 74,398 | 15.3 | 1.01 (0.71-1.44), p=0.95 |
| HTN I15 | <10 | N/A | 8,676 | 2.1 | <10 | N/A | 10,052 | 2.1 | 1.16 (0.34-4.00), p=1.00 |
| Seizures G40 | 93 | 39.9 | 29,105 | 6.9 | 115 | 38.7 | 35,860 | 7.4 | 1.00 (0.76-1.31), p=0.98 |
| Depression F33 | 59 | 25.3 | 96,448 | 23.0 | 69 | 23.2 | 118,134 | 24.3 | 1.05 (0.74-1.48), p=0.79 |
| Bipolar F31 | 32 | 13.7 | 72,757 | 17.4 | 45 | 15.2 | 90,538 | 18.6 | 0.88 (0.56-1.39), p=0.60 |
| Schizophrenia  F20 | <10 | N/A | 14,972 | 3.6 | 12 | 4.0 | 19,046 | 3.9 | 0.53 (0.19-1.50), p=0.22 |
| Anxiety F41 | 171 | 73.4 | 250,293 | 59.7 | 199 | 67.0 | 297,352 | 61.2 | 1.02 (0.83-1.25), p=0.84 |
| Benign  neoplasm D33 | 13 | 5.6 | 224 | 0.1 | 19 | 6.4 | 285 | 0.1 | 0.87 (0.42-1.80), p=0.71 |
| Neoplasm  D43 | 19 | 8.2 | 63 | 0.0 | 20 | 6.7 | 78 | 0.0 | 1.18 (0.58-2.39), p=0.65 |

Table 1: Demographics and comorbid conditions of PMBT incidence 10-years following buprenorphine or naltrexone exposure.
