## Supplemental Table 1 for "Ten-Year Risk of Primary Malignant Brain Tumors After Buprenorphine vs Naltrexone Exposure: A Retrospective Cohort Study Using the Epic Cosmos Database"

|  | **None** | | **None** | |
| --- | --- | --- | --- | --- |
|  | **PMBT (N=1,365)** | | **None (N=2,290,229)** | |
|  | Count | **%** | Count | **%** |
| **Age group** |  |  |  |  |
| 18-45 | 321 | 23.5 | 780,119 | 34.1 |
| 45-65 | 591 | 43.3 | 878,660 | 38.4 |
| ≥65 | 453 | 33.2 | 631,450 | 27.6 |
| **Sex** |  |  |  |  |
| Female | 662 | 48.5 | 968,876 | 42.3 |
| Male | 633 | 46.4 | 923,752 | 40.3 |
| **Race** |  |  |  |  |
| White | 1084 | 79.4 | 1,502,948 | 65.6 |
| Black | 168 | 12.3 | 299,984 | 13.1 |
| Asian | 19 | 1.4 | 22,115 | 1.0 |
| Other | 156 | 11.4 | 237,603 | 10.4 |
| **BMI** |  |  |  |  |
| <18.5 | 63 | 4.6 | 205,389 | 9.0 |
| 18.5-24.9 | 395 | 28.9 | 959,624 | 41.9 |
| 25-29.9 | 464 | 34.0 | 1,122,092 | 49.0 |
| ≥30 | 561 | 41.1 | 1,053,861 | 46.0 |
| **Smoking** |  |  |  |  |
| Never | 470 | 34.4 | 534,607 | 23.3 |
| Current | 335 | 24.5 | 702,858 | 30.7 |
| Former | 451 | 33.0 | 584,607 | 25.5 |
| **Underlying conditions** |  |  |  |  |
| AUD F10 | 199 | 14.6 | 331,491 | 14.5 |
| Other SUD F12-19 | 711 | 52.1 | 1,144,209 | 50.0 |
| Hepatitis B19 | 89 | 6.5 | 148,127 | 6.5 |
| T2DM E11 | 435 | 31.9 | 516,307 | 22.5 |
| HTN I15 | 66 | 4.8 | 68,907 | 3.0 |
| Seizures G40 | 396 | 29.0 | 121,324 | 5.3 |
| Depression F33 | 284 | 20.8 | 354,451 | 15.5 |
| Bipolar F31 | 115 | 8.4 | 208,440 | 9.1 |
| Schizophrenia F20 | 27 | 2.0 | 57,284 | 2.5 |
| Anxiety F41 | 817 | 59.9 | 975,007 | 42.6 |
| Benign neoplasm D33 | 102 | 7.5 | 1,495 | 0.1 |
| Neoplasms D43 | 82 | 6.0 | 377 | 0.0 |

Supplementary Table 1: Demographics and comorbid conditions of PMBT incidence 10-years following non-exposure.
